# Automated scout-image-based estimation of contrast agent dosing: a deep learning approach

**DOI:** 10.1101/2025.05.11.25327410

**Authors:** Robin Tibor Schirrmeister, Laetitia Taleb, Paul Friemel, Marco Reisert, Fabian Bamberg, Jakob Weiß, Alexander Rau

## Abstract

We developed and tested a deep-learning-based algorithm for the approximation of contrast agent dosage based on computed tomography (CT) scout images. We prospectively enrolled 817 patients undergoing clinically indicated CT imaging, predominantly of the thorax and/or abdomen. Patient weight was collected by study staff prior to the examination 1) with a weight scale and 2) as self-reported. Based on the scout images, we developed an EfficientNet convolutional neural network pipeline to estimate the optimal contrast agent dose based on patient weight and provide a browser-based user interface as a versatile open-source tool to account for different contrast agent compounds. We additionally analyzed the body-weight-informative CT features by synthesizing representative examples for different weights using in-context learning and dataset distillation. The cohort consisted of 533 thoracic, 70 abdominal and 229 thoracic-abdominal CT scout scans. Self-reported patient weight was statistically significantly lower than manual measurements (75.13 kg vs. 77.06 kg; p < 10^−5^, Wilcoxon signed-rank test). Our pipeline predicted patient weight with a mean absolute error of 3.90 ± 0.20 kg (corresponding to a roughly 4.48 - 11.70 ml difference in contrast agent depending on the agent) in 5-fold cross-validation and is publicly available at https://tinyurl.com/ct-scout-weight. Interpretability analysis revealed that both larger anatomical shape and higher overall attenuation were predictive of body weight. Our open-source deep learning pipeline allows for the automatic estimation of accurate contrast agent dosing based on scout images in routine CT imaging studies. This approach has the potential to streamline contrast agent dosing workflows, improve efficiency, and enhance patient safety by providing quick and accurate weight estimates without additional measurements or reliance on potentially outdated records. The model’s performance may vary depending on patient positioning and scout image quality and the approach requires validation on larger patient cohorts and other clinical centers.

**Author Summary:** Automation of medical workflows using AI has the potential to increase reproducibility while saving costs and time. Here, we investigated automating the estimation of the required contrast agent dosage for CT examinations. We trained a deep neural network to predict the body weight from the initial 2D CT Scout images that are required prior to the actual CT examination. The predicted weight is then converted to a contrast agent dosage based on contrast-agent-specific conversion factors. To facilitate application in clinical routine, we developed a user-friendly browser-based user interface that allows clinicians to select a contrast agent or input a custom conversion factor to receive dosage suggestions, with local data processing in the browser. We also investigate what image characteristics predict body weight and find plausible relationships such as higher attenuation and larger anatomical shapes correlating with higher body weights. Our work goes beyond prior work by implementing a single model for a variety of anatomical regions, providing an accessible user interface and investigating the predictive characteristics of the images.

## Introduction

The administration of contrast agents is essential in various computed tomography (CT) examinations as this substantially improves the delineation of distinct anatomical structures, and their enhancement patterns facilitate the detection of pathologies [1]. Moreover, correct contrast agent dosing allows for reproducible imaging required for diagnosis and longitudinal follow-up [1]. Therefore, this constitutes an important step in the patients’ diagnostic journey. However, optimal dosage requires human interaction as optimal imaging results need patient-specific dosing of the contrast agent based on body weight, typically obtained through patient self-reporting or technician estimation, both of which may introduce bias and inaccuracies [2], [3]. Direct weight measurements using calibrated scales might provide the correct current weight, reduce bias and facilitate precise dosing but at the cost of substantially increasing the workload. Thus, it is not frequently integrated in the workflow. Deep learning algorithms have the potential to obtain patient-specific information from imaging data. A thus-derived body weight estimation might reduce workload and bias and improve reproducibility when determining optimal contrast agent doses for CT examinations.

Previous work has demonstrated the feasibility of deep-learning-based weight prediction from specific anatomical regions in CT scout images, which are required prior to the actual CT examination to estimate radiation dosage and adjust scan range. Studies have used deep neural networks to predict body weight from either thoracic or abdominal CT scout data separately. One study focused on pediatric patients [4] while another examined the general population [5]. However, these studies were limited to analyzing thoracic or abdominal scans in isolation. Moreover, the applicability to clinical routine workflows is lacking as no user interface for contrast agent dosing was proposed. In addition, no in-depth analysis of the underlying predictive features was conducted, which may help to improve acceptability by users and patients alike and identify challenging constellations that might require human interaction.

To address these limitations, we developed a deep-learning pipeline that estimates contrast agent dosage from CT scout images of a diverse, real-world dataset and provides an easy-to-use browser-based interface. For this, we prospectively collected a clinical dataset of CT scout images and weight-scale measurements, and trained a deep network to predict body weight on these images, which are already collected during regular clinical CT examinations. We implemented this model in a browser-based interface that automatically estimates the optimal dosage for various contrast agents and can be easily integrated into clinical workflows. Additionally, we employed deep network interpretability methods to analyze the predictive features by distilling synthetic representative example training images for different body weights.

## Results and Discussion

### Participants

The cohort consisted of 533 thoracic, 70 abdominal and 229 thoracic-abdominal CT scout scans. Patients were between 18 and 92 years old, 403 male (48.8%) and 429 female (51.6%). The patients had a large variety of body weights as shown in Figure 1. Thirthy-eight patients had to be excluded due to corrupted image quality of the scout, no weight could be reported by the patient, or if scale-based weighting was not feasible, e.g. due to immobility.

**Figure 1:**
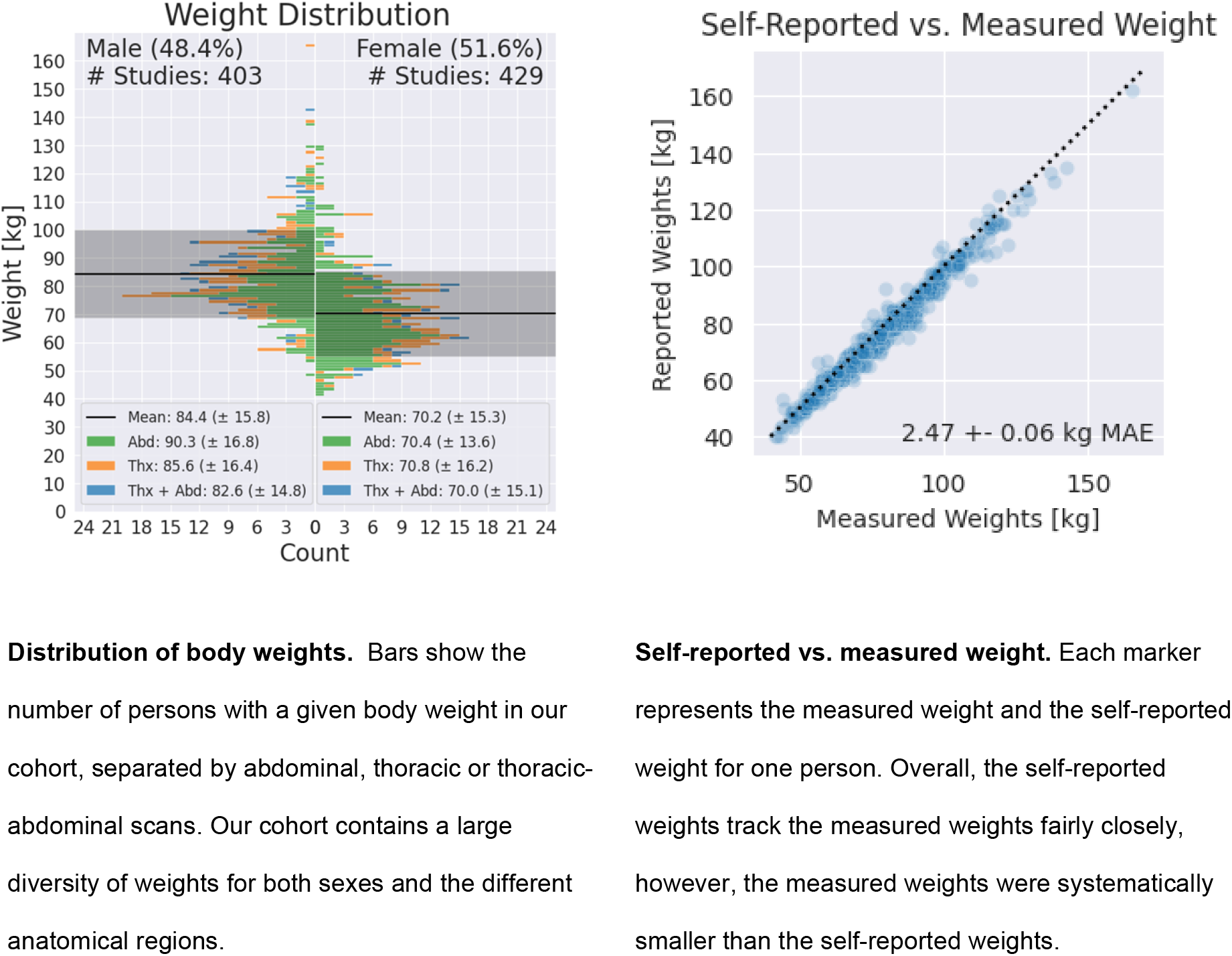

The reported weights are reasonably accurate, yet systematically underestimate the measured weights. After removing seven cases with apparent data entry errors (such as recorded height instead of weight), we found that self-reported weights were significantly lower than manual measurements (75.13 kg vs. 77.06 kg; p < 10^−5^, Wilcoxon signed-rank test). With a mean absolute difference to the measured weights of 2.47 ± 0.06 kg across all studies, these self-reported weights provided a strong baseline for evaluating the prediction differences of our automated pipeline and the measured weights.

### Model testing

In the whole dataset, using 5-fold cross-validation, we found an overall high performance of automated weight prediction with a mean deviation of 3.90 ± 0.20 kg (mean and standard deviation across the 5 folds). Higher performance was noted in scans that encompassed the thorax and abdomen (3.07 ± 0.26 kg), while slightly lower performance was noted for scans of either the abdomen (4.49 ± 0.30 kg) or the thorax (5.67 ± 0.38 kg). The superior performance on thoracic-abdominal scans can likely be attributed to the larger body area providing more weight-relevant information. Notably, the model performed well on abdominal scans despite their limited representation in the dataset (<10% of total data). Exemplary cases of well-predicted as well as over- and underestimated weight are shown in Figure 3.

Analysis of prediction errors revealed no clear patterns across anatomical regions, with well-predicted, underestimated, and overestimated cases distributed across all scan types. However, as shown in Figure 2, the model tends to underestimate larger weights more than smaller weights—a common phenomenon in regression tasks where predictive uncertainty drives predictions toward the dataset mean to minimize expected loss.

**Figure 2:**
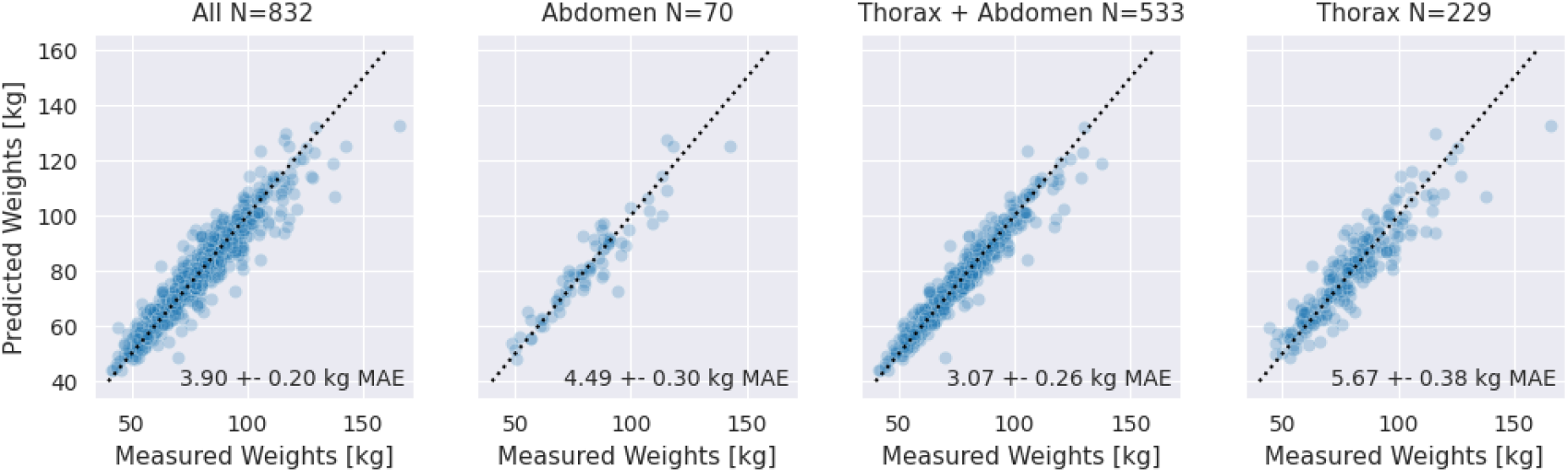
Model performance in CT-scout-based weight prediction. Each marker represents the measured and predicted weight for one person. Predictions for all regions have a lower mean absolute error (MAE) than 5.5, thoracic-abdominal scans are predicted most accurately.

**Figure 3:**
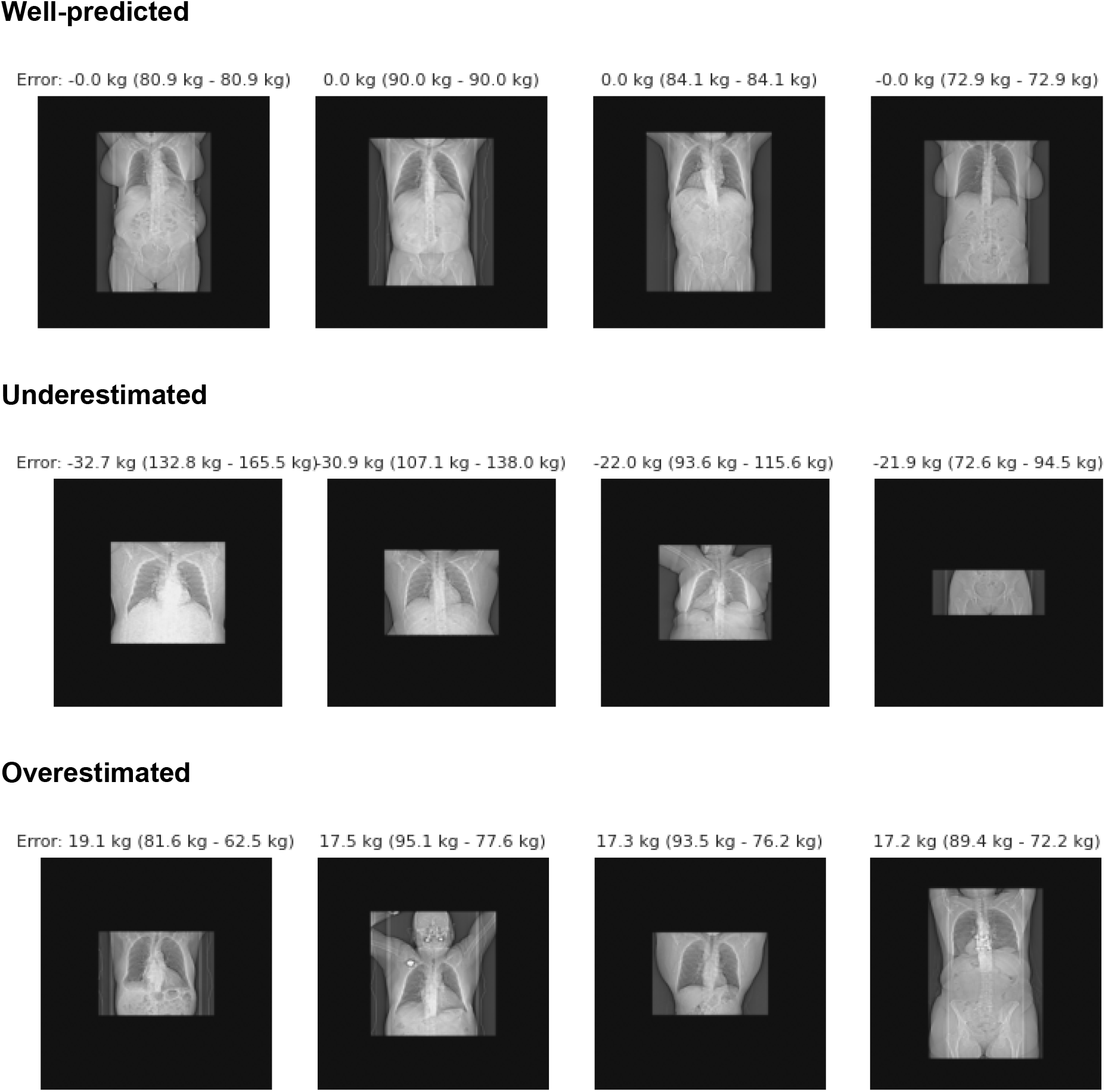
Examples of well-predicted, underestimated, and overestimated cases. Scans from a variety of regions appear in the well-predicted, underestimated, and overestimated categories, with no obvious patterns except for lower weights being more overestimated and higher weights more underestimated.

### Model interpretability

Visual analysis of the original CT scout scans downsampled to 64×64 and sorted by measured body weight revealed mixed patterns as seen in Figure 4. While patients with higher body weight generally showed larger attenuations and anatomical shapes as expected, substantial variation existed among scans from patients of similar weights. This variability makes it challenging to identify which specific features are sufficient for robust predictions.

**Figure 4:**
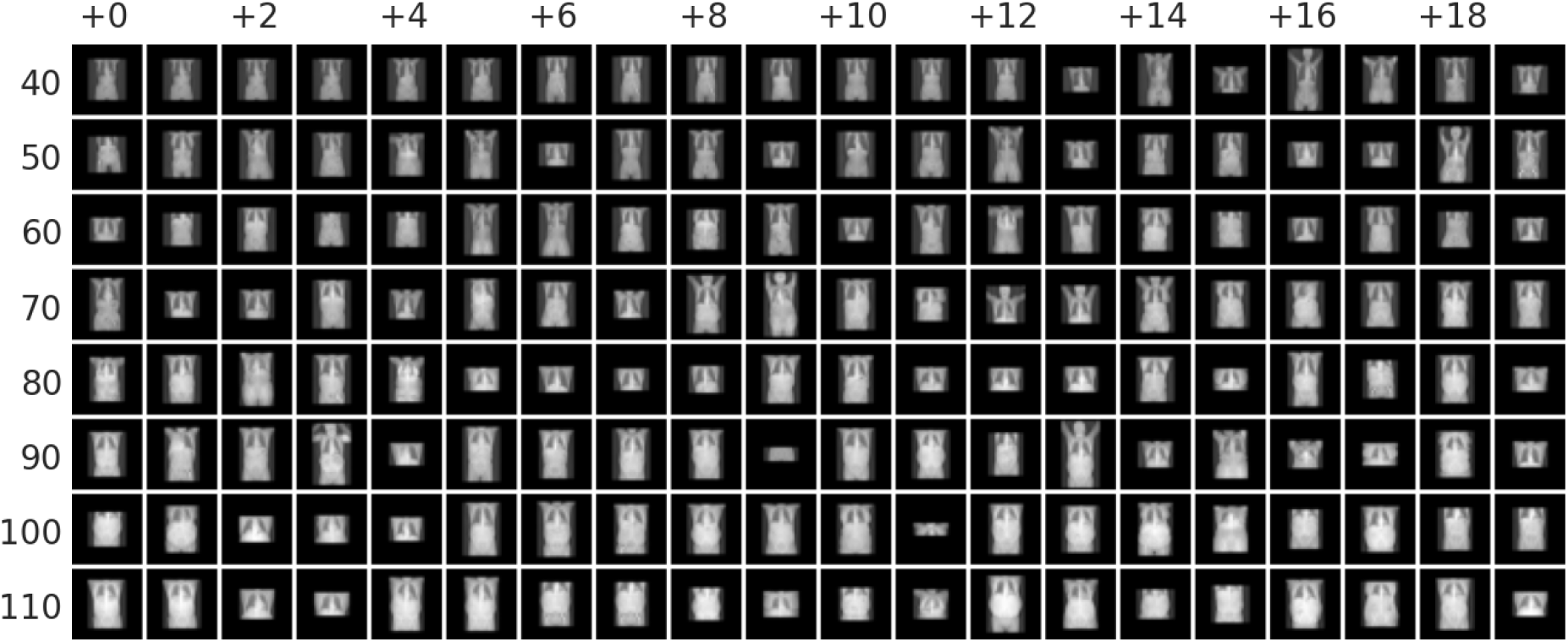
Selected CT Scout images from lower to higher weights in kg. Input images are shown as they are supplied to the deep network during training. A large diversity of shapes is visible, with images with larger body weight having larger shapes and more attenuation.

We used in-context learning combined with dataset distillation to generate a synthetic training dataset that yields high accuracy when used for learning the body weight prediction. The synthetic distilled training dataset in Figure 5 shows a more consistent pattern from lower attenuation and smaller anatomical shape to higher attenuation and larger anatomical shape with increasing body weight. Using in-context learning with this distilled dataset still retains an average MAE below 5 kg, indicating that the contained information is enough to explain a substantial amount of the predictive features. Overall, this suggests that the expected relationship of attenuation and anatomical size with body weight contains sufficient information for accurate predictions even after substantial downsampling of the data. The remaining gap in the results of the network trained on the full-resolution data may either be explicable purely by the larger resolution or by additional learned features beyond those visible in the distilled dataset.

**Figure 5:**
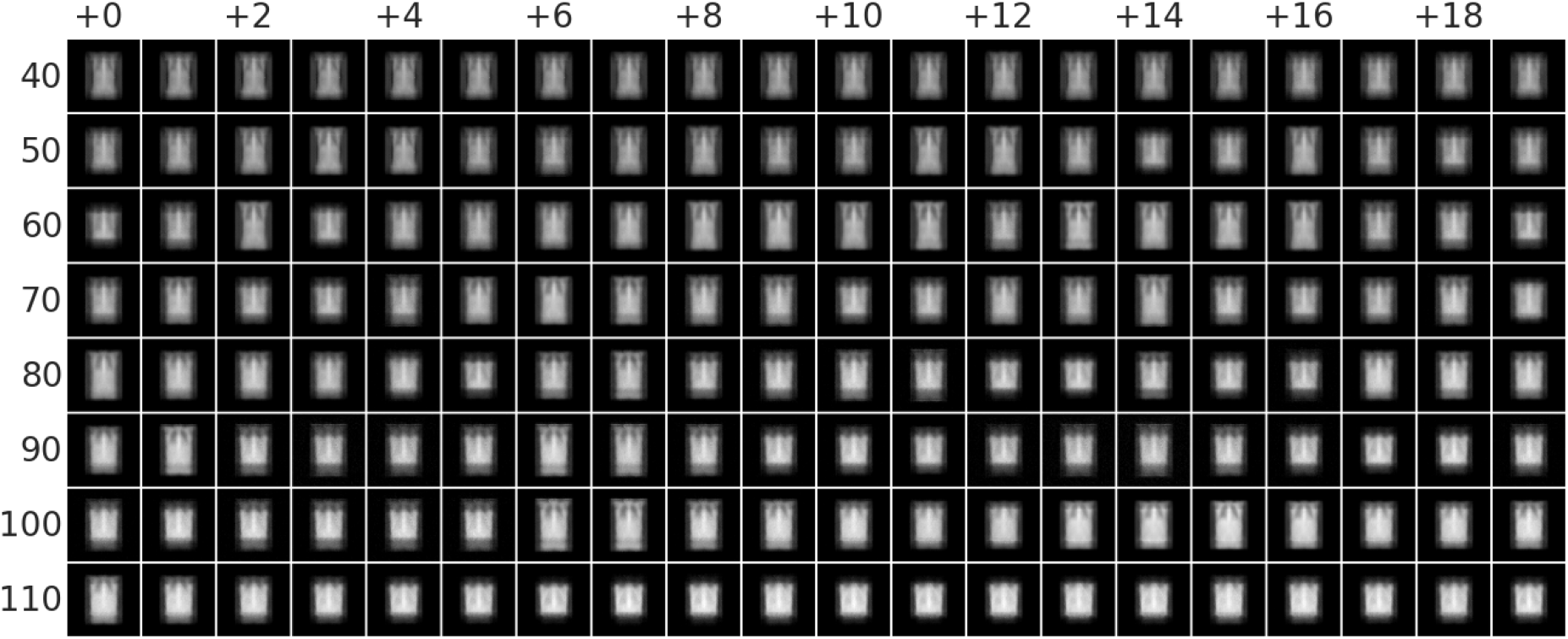
Synthetic distilled dataset from lower to higher weights in kg. Dataset was synthesized to perform well as an in-context training set and is sufficient to reach below 5 kg mean absolute error. Note the smooth progression from smaller anatomical shapes and less attenuation to larger shapes and stronger attenuation with increasing body weight.

### Open-source web user interface

We developed a browser-based user interface that enables CT scout image upload and provides contrast-agent-specific dosage estimates using the hereby developed model. Users can input CT scout scans as NIFTI files and select contrast agents from a dropdown menu, with dosage suggestions calculated using either manufacturer-provided or custom conversion factors as can be seen in Figure 6. The interface runs locally using ONNX Runtime [6], ensuring data privacy while delivering weight predictions in approximately one second on standard hardware (Intel Xeon CPU with 6 Cores at 3.5 GHz, no GPU). The tool is publicly available at https://tinyurl.com/ct-scout-weight.

**Figure 6.**
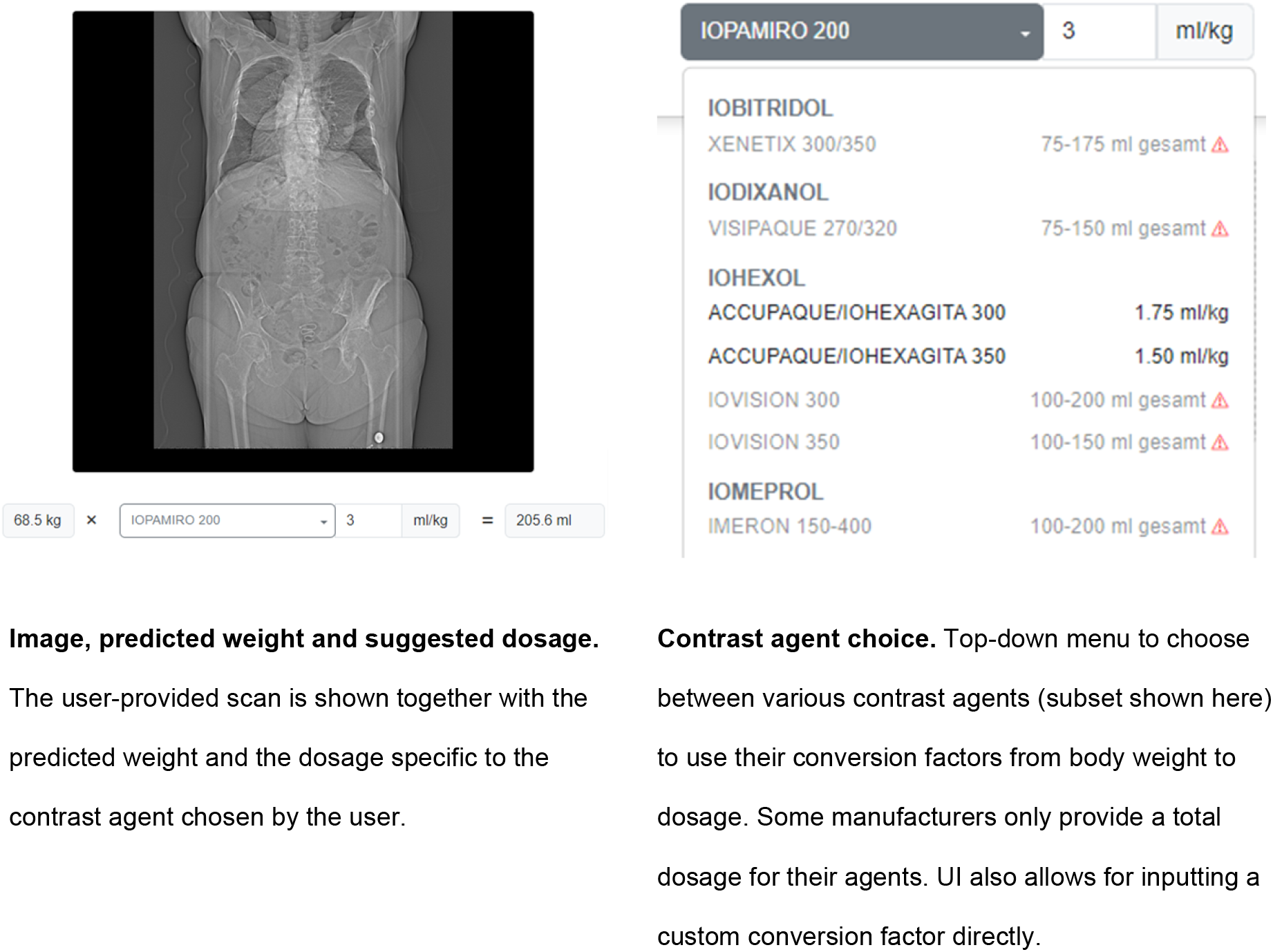
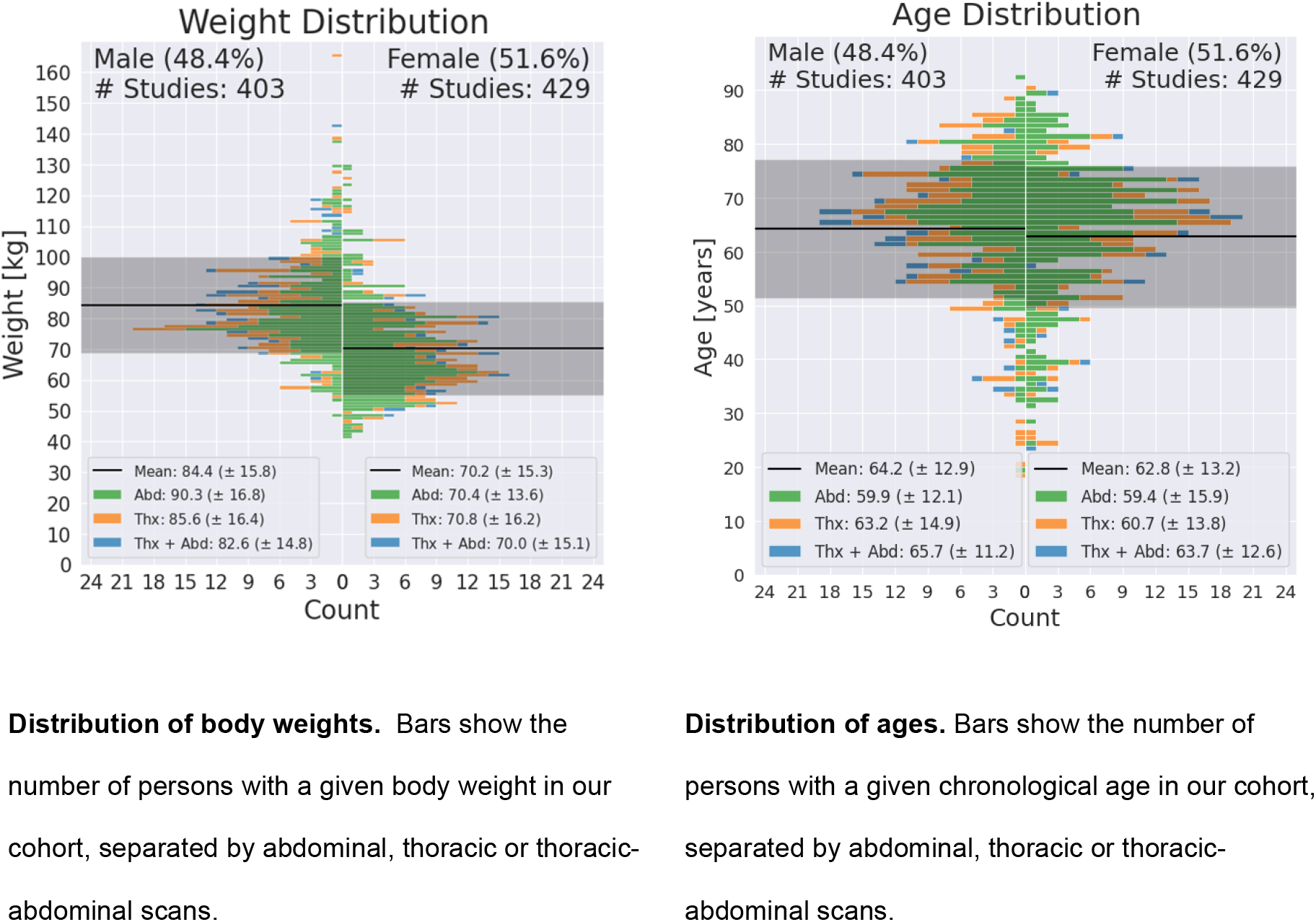
Distribution plots for weights and ages. Our cohort contains a large diversity of weights and ages for both sexes and the different anatomical regions.

## Conclusion

Our study demonstrates a deep learning-based workflow for estimating contrast agent dosage from CT scout scans applicable to clinical routine. With a mean absolute error below 4 kg in weight estimation and interpretability analysis confirming plausible trustworthy predictive features, the approach shows substantial potential for clinical utility. The browser-based interface enables practical implementation in hospital settings while maintaining data privacy. Future work should validate this approach across larger cohorts and multiple clinical centers.

## Materials and Methods

### Dataset

In this prospective study, we included consecutive patients who underwent clinically indicated contrast-enhanced CT imaging of the thorax and/or abdomen at our tertiary referral center. All patients were scanned on either a NAEOTOM.alpha or Somatom.Edge (Siemens Healthineers, Erlangen, Germany) using routine protocols. Scout data were obtained without any dose modulation and fixed 100 kV and 55 mAs (NAEOTOM.alpha) or 75 mAs (Somatom.Edge), respectively, with top tube position. Body weight was both obtained via patients’ self reports and weight-scale measurements by study staff. Age and sex were obtained from clinical records.

Patients were excluded if they were below 18 years of age or due to corrupted image quality of the scout, no weight could be reported by the patient, or if scale-based weighting was not feasible, e.g. due to immobility.

Our cohort contains a large diversity of weights and ages for both sexes and the different anatomical regions, see Figure 6.

This study was approved by the Institutional Review Board (Ethic Committee Freiburg 21– 2469). All participants provided informed consent.

### Model development and testing

#### Development

For weight prediction from CT scout scans, we employed the widely-used EfficientNet architecture [7], specifically its EfficientNet-B1 variant, implemented through the MONAI library [8]. This architecture has demonstrated success in preliminary experiments and has previously been used for CT scout-based weight prediction [4]. We optimized the network parameters using AdamW with an L1-Loss function, using a learning rate of 1.5 × 10^−4^ and weight decay of 10^−5^. For data augmentation, we implemented a subset of techniques from nnUNet, namely random cropping, low-resolution simulation, Gaussian noise addition, Gaussian blurring, and random gamma transformation.

#### Testing

The model was tested via 5-fold cross-validation, with the training folds always containing 80% and the test folds containing the remaining 20% of the full dataset. We report overall results as well as results grouped by the different anatomical regions.

### Model interpretability

To gain insight and interpretability of the model, we analyzed predictive features for body weight estimation, by combining a variational autoencoder with in-context learning. In-context learning, for example with the TabPFN network [9], uses an entire training dataset as context to predict test data without any training or finetuning of the network parameters. Crucially, this approach enables the distillation of a smaller synthetic training dataset that performs well as context to predict the original dataset. For a general explanation of this context-tuning method, see [10]. The synthetic dataset provides a compact visual representation of the essential predictive features. In order to be able to apply TabPFN, we downsampled the images to 32×32 resolution and subsequently trained a variational autoencoder to further compress the images into a latent 16-dimensional representation. We then visualized the entire synthetic training dataset to identify visual patterns potentially predictive of body weight.

### Statistical analysis

We evaluated our results using two statistical approaches: a Wilcoxon signed-rank test to compare reported versus measured weights, and cross-validation to assess prediction accuracy. The Wilcoxon test determined if the differences between reported and measured weights were statistically significant. From the 5-fold cross-validation, we calculated both the average mean absolute deviation across folds and its standard error to quantify prediction performance.

## Data Availability

The UI is publicly available at as stated in the manuscript https://tinyurl.com/ct-scout-weight UI and training code is available under https://osf.io/y2pzc/?view_only=cfdd74947e4845528d7b66ca58b01f9a Training data is available upon reasonable research request to comply with our data regulations regarding clinical, sensitive data.

https://nora-imaging.org/ct-scout-weight/

## References

[1] K. T. Bae, „Intravenous contrast medium administration and scan timing at CT: considerations and approaches”, Radiology, Bd. 256, Nr. 1, S. 32–61, Juli 2010, doi: 10.1148/radiol.10090908.

[2] J. Boos u. a., „Does body mass index outperform body weight as a surrogate parameter in the calculation of size-specific dose estimates in adult body CT?”, Br. J. Radiol., Bd. 89, Nr. 1059, S. 20150734, 2016, doi: 10.1259/bjr.20150734.

[3] M. Fukunaga, K. Matsubara, S. Ichikawa, H. Mitsui, H. Yamamoto, und T. Miyati, „CT dose management of adult patients with unknown body weight using an effective diameter”, Eur. J. Radiol., Bd. 135, S. 109483, Feb. 2021, doi: 10.1016/j.ejrad.2020.109483.

[4] A. Demircioğlu, A. S. Quinsten, L. Umutlu, M. Forsting, K. Nassenstein, und D. Bos, „Determining body height and weight from thoracic and abdominal CT localizers in pediatric and young adult patients using deep learning”, Sci. Rep., Bd. 13, Nr. 1, S. 19010, Nov. 2023, doi: 10.1038/s41598-023-46080-5.

[5] S. Ichikawa, M. Hamada, und H. Sugimori, „A deep-learning method using computed tomography scout images for estimating patient body weight”, Sci. Rep., Bd. 11, Nr. 1, S. 15627, Aug. 2021, doi: 10.1038/s41598-021-95170-9.

[6] ONNX Runtime developers, ONNX Runtime. (November 2018). C++. Zugegriffen: 26. März 2025. [Online]. Verfügbar unter: https://github.com/microsoft/onnxruntime

[7] M. Tan und Q. Le, „Efficientnet: Rethinking model scaling for convolutional neural networks”, in International conference on machine learning, PMLR, 2019, S. 6105–6114.

[8] M. J. Cardoso u. a., MONAI: An open-source framework for deep learning in healthcare. (November 2022). Python. doi: 10.48550/arXiv.2211.02701.

[9] N. Hollmann u. a., „Accurate predictions on small data with a tabular foundation model”, Nature, Bd. 637, Nr. 8045, S. 319–326, Jan. 2025, doi: 10.1038/s41586-024-08328-6.

[10] B. Feuer u. a., „TuneTables: Context Optimization for Scalable Prior-Data Fitted Networks”, gehalten auf der The Thirty-eighth Annual Conference on Neural Information Processing Systems, Nov. 2024. Zugegriffen: 12. November 2024. [Online]. Verfügbar unter: https://openreview.net/forum?id=FOfU3qhcIG&referrer=%5Bthe%20profile%20of%20Colin%20White%5D(%2Fprofile%3Fid%3D~Colin_White1)

